# Association between gene expression profiling of skin lesion and autoantibody in patients with systemic sclerosis

**DOI:** 10.1101/2020.04.12.20063131

**Authors:** Jun Inamo

## Abstract

**Objective:** The aim of this study was to investigate relevance between type of autoantibody and gene expression profile in skin lesion of systemic sclerosis (SSc), and identify specifically dysregulated pathways.

**Methods:** Sixty-one patients with SSc from the Genetics versus Environment in Scleroderma Outcome Study cohort and thirty-six healthy controls (HC) are included. Differentially expressed genes (DEGs) were extracted and functional enrichment and pathways analysis were conducted.

**Results:** Compared with HC, lists consisting of 2, 71, 10, 144 and 78 DEGs were created for patients without specific autoantibody, anti-centromere (ACA), anti-U1 RNP (RNP), anti-RNA polymerase III (RNAP) and anti-topoisomerase I (ATA) antibody, respectively. While part of enriched pathways overlapped, distinct pathways were identified except those without specific autoantibody: *keratinocyte differentiation* in ACA, *NF-kB signaling* and *cellular response to transforming growth factor beta stimulus* in RNAP, *interferon alpha/beta signaling* of RNP and *cellular response to stress* in ATA.

**Conclusion:** Pathogenic pathways were identified according to type of autoantibodies by leveraging gene expression data of patients and controls from multi-center cohort. The current study will promote to explore new therapeutic target for SSc.

**Key message:** Distinct pathways are associated with type of autoantibody in skin lesion of systemic sclerosis.

## Introduction

Systemic sclerosis (SSc) is a heterogeneous autoimmune disease characterized by fibrosis and micro-vasculopathy. Compared with limited cutaneous systemic sclerosis (lcSSc), those with diffuse cutaneous subtype of SSc (dcSSc) have with high morbidity and mortality. Progression of skin lesion is associated with subsequent progression of visceral organ lesion and mortality in dcSSc, suggesting critical disorder may be shared among various lesion [1]. Thus, skin biopsy has been conducted to explore the key molecule in the pathogenesis of dcSSc. To date, transcriptome profiling has revealed involvement of fibroinflammatory pathways in aberrant milieu of skin lesion [2,3]. According to gene expression profiles observed in their skin biopsy specimens, there are 3 subgroups of patients with dcSSc; an inflammatory pattern, a proliferative pattern and normal-like pattern [4].

However, while type of autoantibodies is relevant to clinical traits, association between gene expression profile in situ and autoantibodies is still largely unknown. Antinuclear autoantibodies are positive in more than 80% of patients with SSc [5,6]. Anti-centromere (ACA) antibody is associated with lcSSc and negatively associated with interstitial lung disease (ILD) [5,6]. Anti-topoisomerase I (ATA) antibody, which is almost specific for dcSSc, is a risk factor for ILD [5,6]. Anti-RNA polymerase III (RNAP) antibody is almost specific for dcSSc and associated with renal crisis [7].

Patients with positivity of anti-U1 RNP (RNP) antibody, which is associated with overlapping features of multiple connective diseases including SSc, develop musculoskeletal involvement earlier and more frequently than other type of SSc [8]. On the other hand, less than 10% of SSc patients are seronegative [9]. These patients are common in dcSSc and characterized by less vasculopathy such as pulmonary hypertension, digital ulcers and fewer telangiectasias, and a greater proportion of males and gastrointestinal involvement [10].

The aim of this study was to investigate relevance between type of autoantibody and gene expression profile in skin lesion, and identify specifically dysregulated pathways.

## Materials and Methods

### Patients and control subjects

The Genetics versus Environment in Scleroderma Outcome Study (GENISOS) is prospective cohort with collaboration between the University of Texas Health Science Center at Houston, the University of Texas Medical Branch at Galveston and the University of Texas Health Science Center at San Antonio [11]. All patients fulfilled the criteria for SSc according to American College of Rheumatology/European League Against Rheumatism classification criteria [12]. Type of SSc, dcSSc or lcSSc, was defined based on the extent of cutaneous involvement [13]. The modified Rodnan skin thickness score [14] was used to assess skin thickness. The presence of interstitial lung disease was defined by high-resolution computed tomography findings and decreased forced vital capacity less than 70% predicted.

### Expression profiling

The methods of skin biopsy and gene expression measurement were described in the previous report [2]. Briefly, after RNA was extracted from skin specimen, global gene expression was measured using Illumina HumanHT-12bead arrays. All microarray experiments were performed in single batch. Raw expression dataset is available in public GEO (accession number GSE58095). Data were normalized according to the median method and transformed to z-score. For quality control, genes whose log intensity variance was in the bottom 75th percentile and whose expression level were in the bottom 20th percentile were filtered out. Finally, 9456 transcripts met this criterion. The extraction of differentially expressed genes (DEGs) was conducted based on the empirical Bayes method in the R/Bioconductor limma package. DEGs were extracted by setting the criterion for statistical significance as an adjusted P value by the Benjamini-Hochberg procedure < 0.05 and an absolute fold change > 1.5. Functional enrichment and pathways analysis were conducted using Gene Ontology terms, Kyoto Encyclopedia of Genes and Genomes pathway, Hall mark gene sets, Canonical Pathways and Reactome Gene Sets. The interactive visualization was generated by Metascape (https://metascape.org/gp/index.html). In hierarchical clustering, genes were divided into clusters by k-means method.

### Statistics

Continuous data were presented as the median and range or as a number with a percentage value, as appropriate. The Wilcoxon rank sum test was used to examine the differences between continuous variables. Fisher’s exact test was used to compare proportions in categorical data between groups. All statistical analyses without transcriptome analyses were performed with R (R Foundation for Statistical Computing, Vienna, Austria).

## Results

Totally, 61 patients with SSc from GENISOS cohort and 36 healthy controls (HC) were included (Table 1). Patients with positivity of RNP were significantly younger, and those with RNAP and ATA had high the modified Rodnan skin thickness score at biopsy.

**Table 1.**
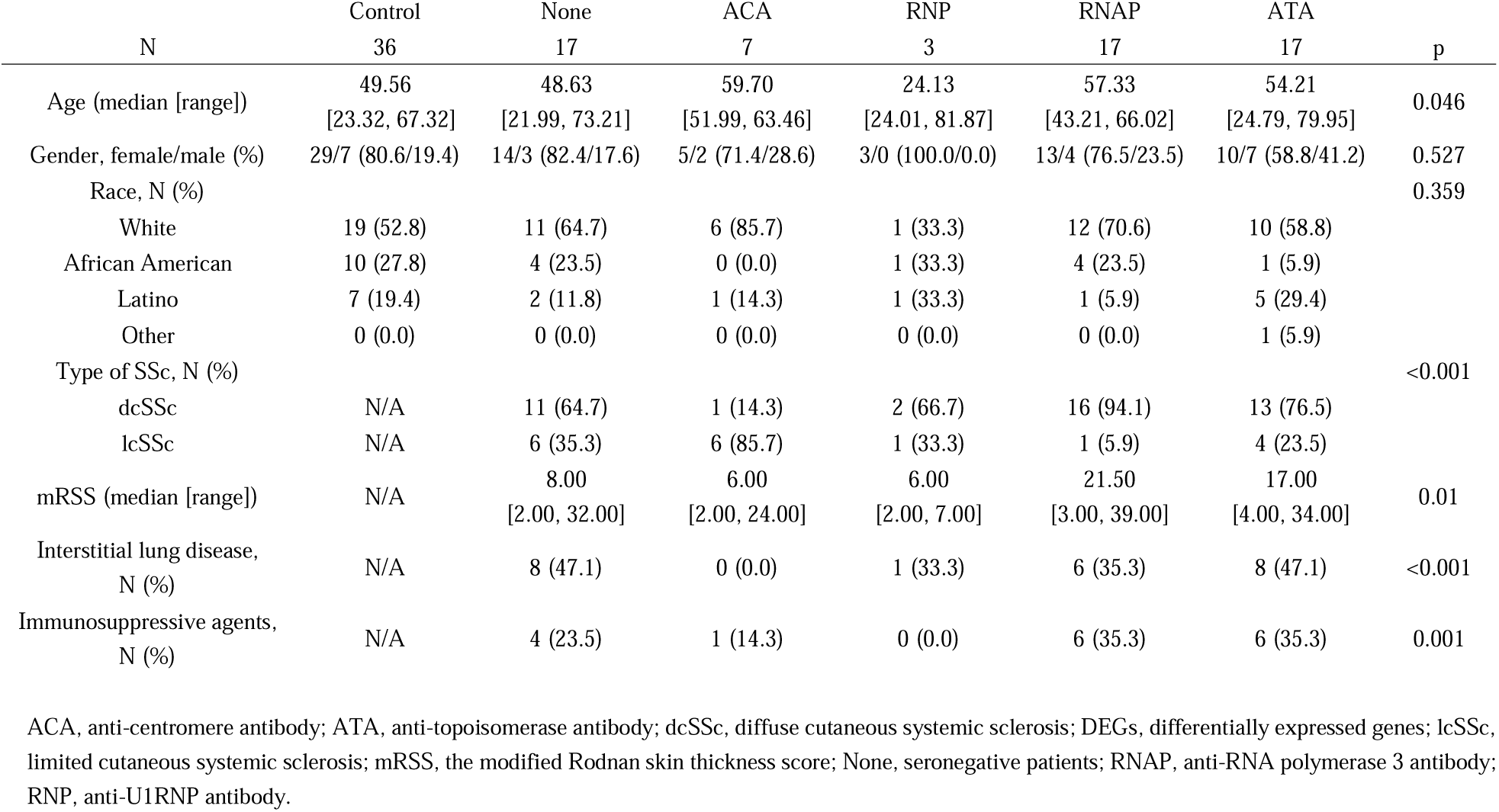
Demographic and clinical characteristics of subjects at skin biopsy.

| N | Control<br>36 | None<br>17 | ACA<br>7 | RNP<br>3 | RNAP<br>17 | ATA<br>17 | p |
| --- | --- | --- | --- | --- | --- | --- | --- |
| Age (median [range]) | 49.56<br>[23.32, 67.32] | 48.63<br>[21.99, 73.21] | 59.70<br>[51.99, 63.46] | 24.13<br>[24.01, 81.87] | 57.33<br>[43.21, 66.02] | 54.21<br>[24.79, 79.95] | 0.046 |
| Gender, female/male (%) | 29/7 (80.6/19.4) | 14/3 (82.4/17.6) | 5/2 (71.4/28.6) | 3/0 (100.0/0.0) | 13/4 (76.5/23.5) | 10/7 (58.8/41.2) | 0.527 |
| Race, N (%) |  |  |  |  |  |  | 0.359 |
| White | 19 (52.8) | 11 (64.7) | 6 (85.7) | 1 (33.3) | 12 (70.6) | 10 (58.8) |  |
| African American | 10 (27.8) | 4 (23.5) | 0 (0.0) | 1 (33.3) | 4 (23.5) | 1 (5.9) |  |
| Latino | 7 (19.4) | 2 (11.8) | 1 (14.3) | 1 (33.3) | 1 (5.9) | 5 (29.4) |  |
| Other | 0 (0.0) | 0 (0.0) | 0 (0.0) | 0 (0.0) | 0 (0.0) | 1 (5.9) |  |
| Type of SSc, N (%) |  |  |  |  |  |  | <0.001 |
| dcSSc | N/A | 11 (64.7) | 1 (14.3) | 2 (66.7) | 16 (94.1) | 13 (76.5) |  |
| lcSSc | N/A | 6 (35.3) | 6 (85.7) | 1 (33.3) | 1 (5.9) | 4 (23.5) |  |
| mRSS (median [range]) | N/A | 8.00<br>[2.00, 32.00] | 6.00<br>[2.00, 24.00] | 6.00<br>[2.00, 7.00] | 21.50<br>[3.00, 39.00] | 17.00<br>[4.00, 34.00] | 0.01 |
| Interstitial lung disease,<br>N (%) | N/A | 8 (47.1) | 0 (0.0) | 1 (33.3) | 6 (35.3) | 8 (47.1) | <0.001 |
| Immunosuppressive agents,<br>N (%) | N/A | 4 (23.5) | 1 (14.3) | 0 (0.0) | 6 (35.3) | 6 (35.3) | 0.001 |

ACA, anti-centromere antibody; ATA, anti-topoisomerase antibody; dcSSc, diffuse cutaneous systemic sclerosis; DEGs, differentially expressed genes; lcSSc, limited cutaneous systemic sclerosis; mRSS, the modified Rodnan skin thickness score; None, seronegative patients; RNAP, anti-RNA polymerase 3 antibody; RNP, anti-U1RNP antibody.

First, whole gene expression profiling was described by hierarchical clustering analysis (Figure 1A). Genes were divided to 3 clusters (Supplementary Table 1): those in cluster 2 were specifically upregulated in SSc, while those in cluster 1 and 3 was not different among SSc and HC. Functional enrichment analysis revealed that cluster 1 was characterized by *ERBB2 signaling*, which is involved in pulmonary fibrosis in bleomycin-treated mice [15] (Figure 1B). Genes in cluster 2 were associated with inflammatory pathways such as *neutrophil degranulation, antigen processing and presentation* and *cytokine mediated signaling pathway*. On the other hand, enriched terms by genes in cluster 3 were physiological function, suggesting normal-like signatures. These results were in agreement with previous report [2].

**Figure 1.**
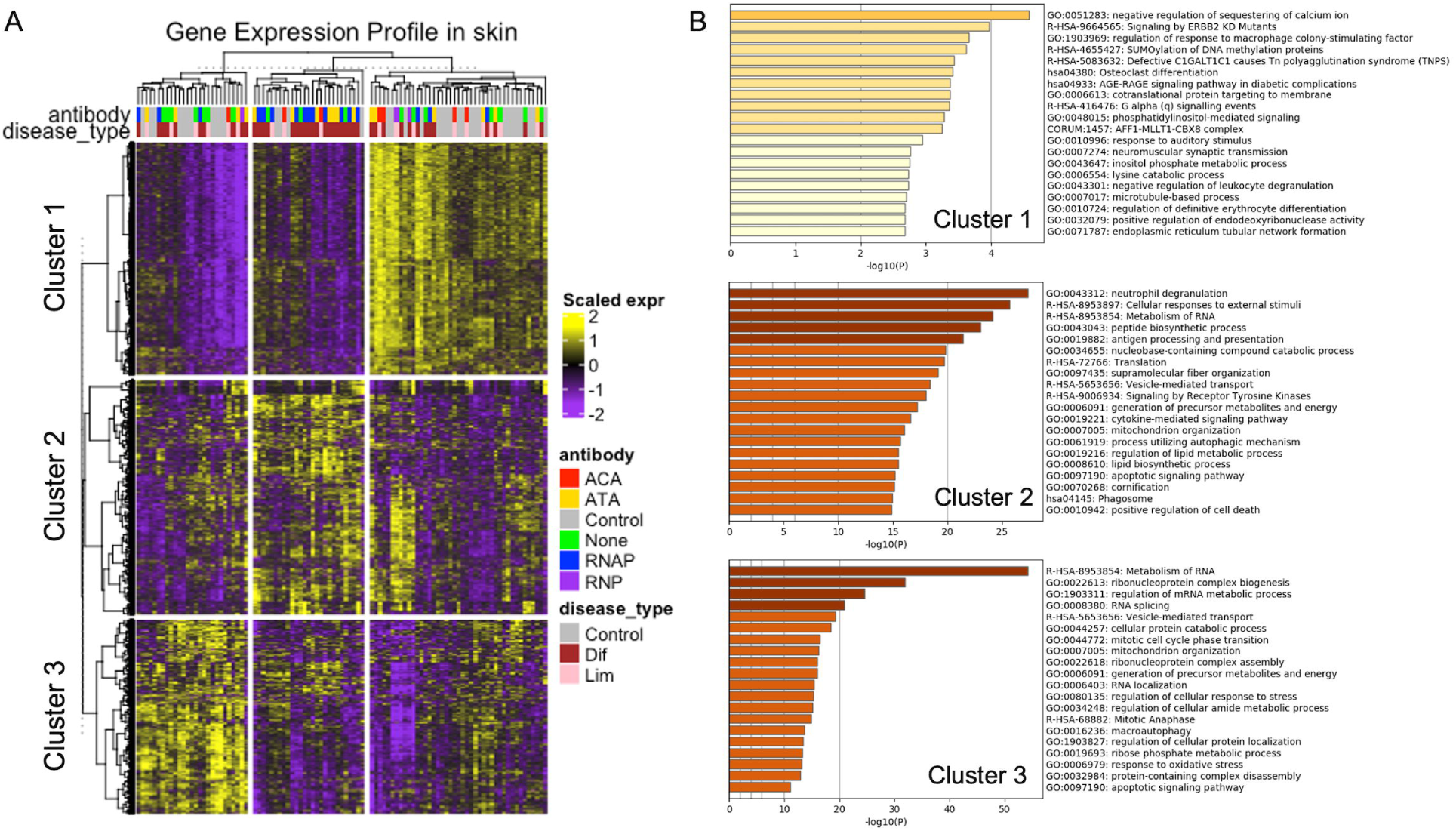
Whole gene expression profiling and functional enrichment analysis. A) Hierarchical clustering analysis. B) Functional enrichment analysis using genes in cluster 1, 2 and 3 identified in hierarchical clustering analysis (A). ACA, anti-centromere antibody; ATA, anti-topoisomerase antibody; dcSSc, diffuse cutaneous systemic sclerosis; lcSSc, limited cutaneous systemic sclerosis; None, seronegative patients; RNAP, anti-RNA polymerase 3 antibody; RNP, anti-U1RNP antibody.

Then, gene expression was compared between patients and controls according to type of autoantibody. Lists consisting of 2, 71, 10, 144 and 78 DEGs were created for patients without specific autoantibody (None), ACA, RNP, RNAP and ATA, respectively (Figure 2A-E and Supplementary Table 2). A part of DEGs were overlapped among ACA, RNAP and ATA, suggesting those might share condition of cutaneous lesion to some extent (Figure 2F and G). For example, *ITGB5*, which is a beta subunit of integrin and receptor for fibronectin, was identified as DEGs of ACA, RNAP and ATA. *TAGLN*, which ubiquitously expresses in vascular and visceral smooth muscle and downstream of transforming growth factor-beta (TGF-β)[16], was also included in DEGs of ACA, RNAP and ATA. Indeed, enrichment analysis using upregulated DEGs demonstrated that the several terms, such as *extracellular matrix organization*, were overlapped among them (Figure 3). On the other hand, signature of DEGs of RNP were distinct from other autoantibodies (Figure 2F and G), and *interferon alpha/beta signaling* was upregulated, representing features of overlapping other connective diseases [8]. Regarding to downregulated DEGs, while *eukaryotic translation elongation* was enriched in RNAP and ATA, its relevance fibrotic diseases is unknown (Figure 3).

**Figure 2.**
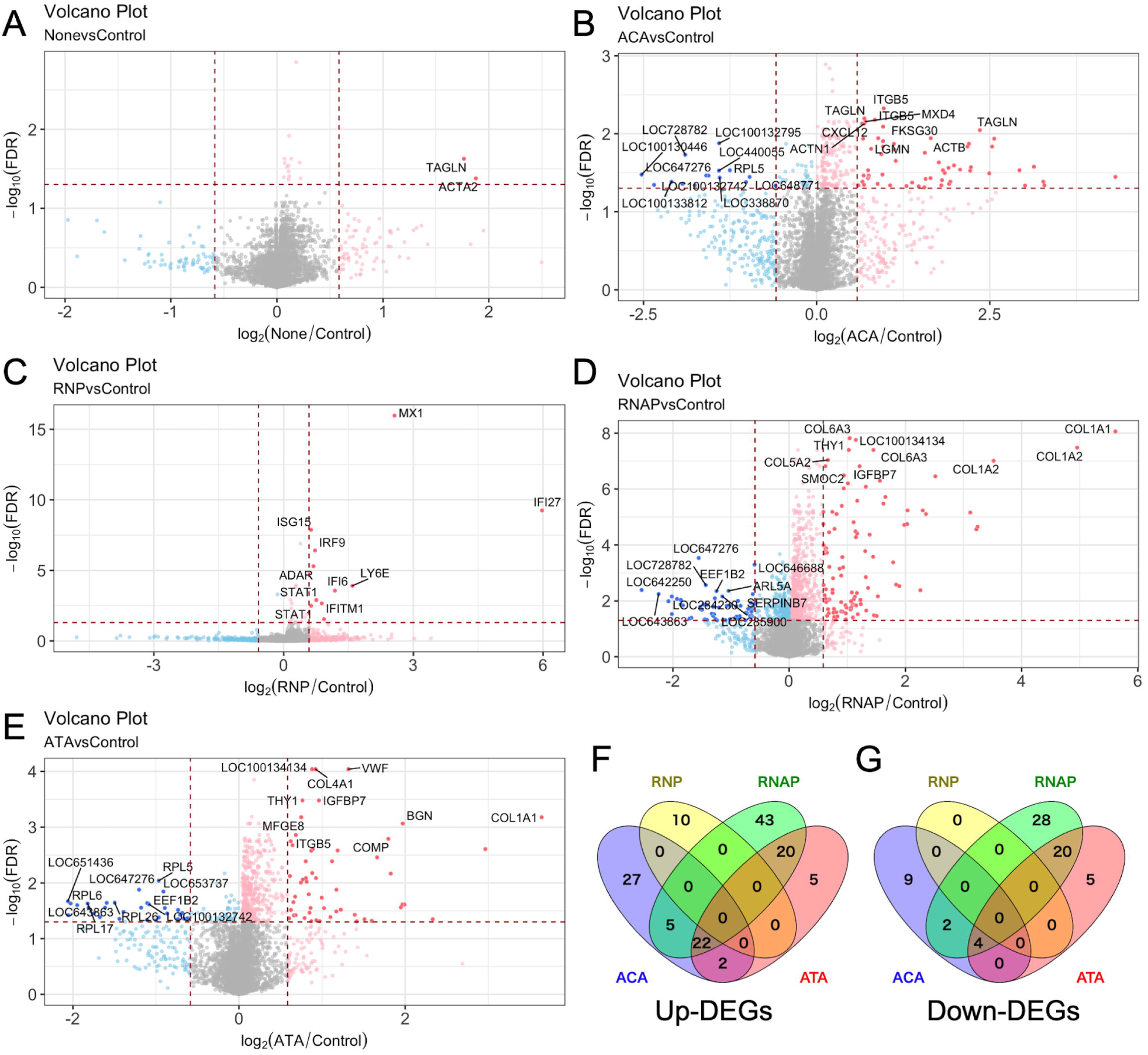
Differentially expressed genes compared with healthy controls. A, B, C, D and E) Volcano plot of None (A), ACA (B), RNP (C), RNAP (D) and ATA (E). Top 10 significantly upregulated and downregulated transcripts are annotated by gene symbol. F and G) Venn diagram of upregulated DEGs (F) and downregulated DEGs (G). ACA, anti-centromere antibody; ATA, anti-topoisomerase antibody; DEGs, differentially expressed genes; None, seronegative patients; RNAP, anti-RNA polymerase 3 antibody; RNP, anti-U1RNP antibody.

**Figure 3.**
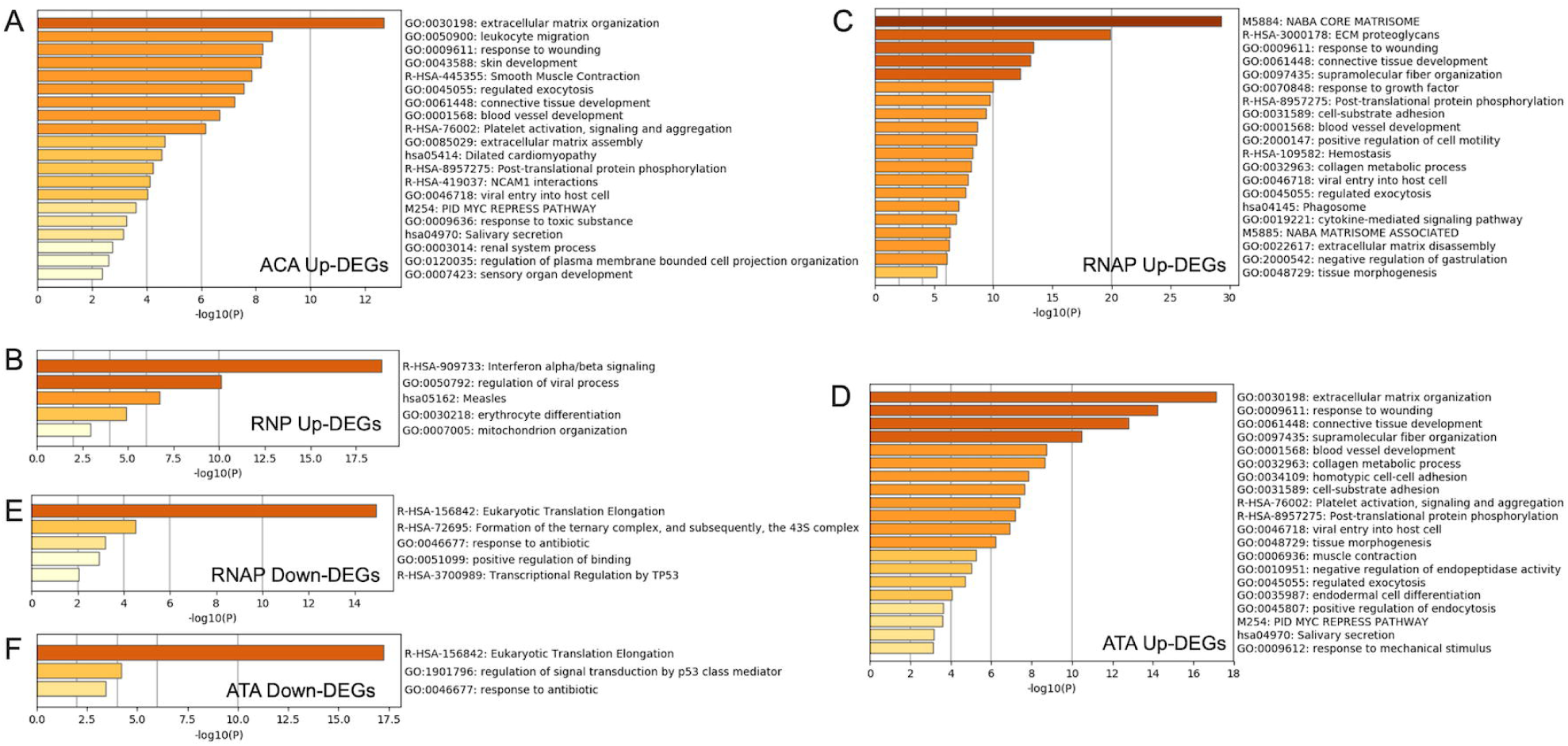
Functional enrichment analysis using differentially expressed genes. A, B, C, D, E and F) Enriched terms using differentially expressed genes in ACA (A), RNP (B), RNAP (C and E) and ATA (D and F). Top 20 significant terms are shown. ACA, anti-centromere antibody; ATA, anti-topoisomerase antibody; DEGs, differentially expressed genes; None, seronegative patients; RNAP, anti-RNA polymerase 3 antibody; RNP, anti-U1RNP antibody.

Finally, to explore distinct pathways characterizing signature of each autoantibody, functional enrichment analysis was conducted using specific DEGs of each autoantibody (Figure 4). Corresponding with the result earlier, *interferon alpha/beta signaling* was upregulated in RNP. Interestingly, while fibrotic pathway such as *keratinocyte differentiation* was enriched in ACA, inflammatory pathways including *NF-kB signaling* also enriched in RNAP. No term was enriched in specifically upregulated DEGs in ATA. According to downregulated DEGs, *ARG1* (Arginase-1) and *CLDN1* (Claudin□1) were relevant to *cellular response to transforming growth factor beta stimulus* in RNAP. Macrophage-specific *ARG1* functions as an inhibitor of inflammation and fibrosis via suppressing T helper 2 (Th2) cytokine [17], suggesting downregulation of *ARG1* might promote fibrosis. *CLDN1*, which is downstream of TGF-β, is required for the normal barrier function of the skin through maintaining tight junctions [18]. As to ATA, *H3-3B, RPL24, RPL31* and *RPL23* were downregulated, implying suppression of *cellular response to stress* (e.g., hypoxia and oxidative stress).

**Figure 4.**
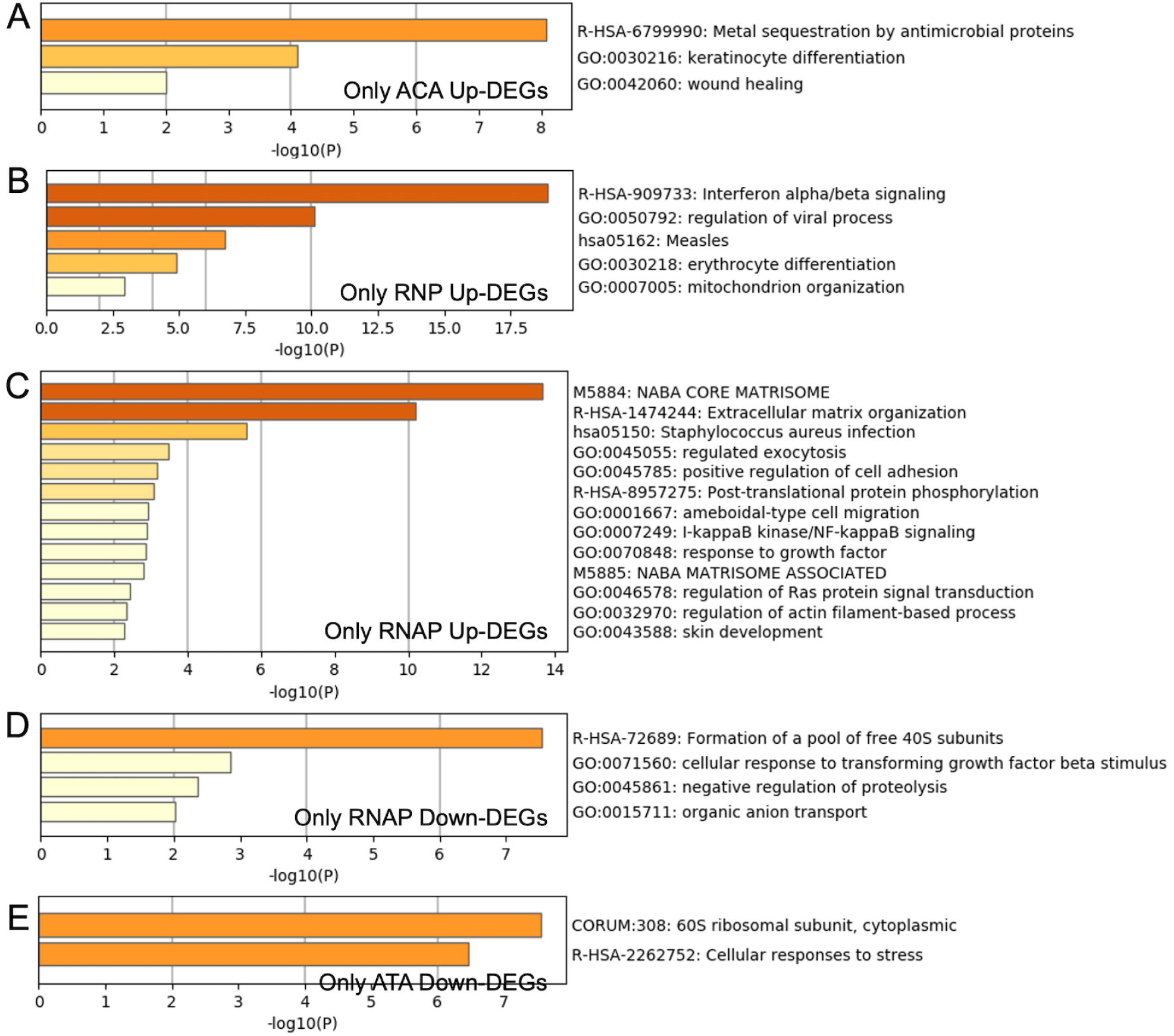
Functional enrichment analysis using differentially expressed genes without overlapping each other. A, B, C, D and E) Enriched terms using differentially expressed genes without overlapping each other in ACA (A), RNP (B), RNAP (C and D) and ATA (E). Top 20 significant terms are shown. ACA, anti-centromere antibody; ATA, anti-topoisomerase antibody; DEGs, differentially expressed genes; None, seronegative patients; RNAP, anti-RNA polymerase 3 antibody; RNP, anti-U1RNP antibody.

## Discussion

The current study reconfirmed that condition of cutaneous lesion were divided into 3 subgroups according to gene expression profiling in skin lesion: fibrosis, inflammation and normal-like signature [2]. In addition, while part of pathogenic pathways overlapped, distinct pathways were identified according to type of autoantibodies, representing characteristics of clinical features autoantibodies of SSc: for example, upregulation of *keratinocyte differentiation* in ACA, *NF-kB signaling* and *cellular response to transforming growth factor beta stimulus* in RNAP, *interferon alpha/beta signaling* of RNP and suppression of *cellular response to stress* in ATA.

The evidence of the mechanisms that lead to chronicity of tissue repair responses in SSc is accumulating. Genome-wide association studies revealed numerous susceptibility loci for SSc and implicated involvement of several inflammatory responses including interferon stimulated genes in the pathogenesis od SSc: *IRF5* (encoding interferon regulatory factor 5), *IRF8* (interferon regulatory factor 8) and *STAT4* (signal transducer and activator of transcription 4) [19]. The current study suggests, although the inflammatory responses in SSc vary across numerous pathways, upregulation of interferon signaling is characteristic especially in SSc overlapping other connective diseases, which is associated with RNP. On the other hand, SSc with RNAP was associated with upregulation of *NF-kB signaling* in skin, corresponding with recent study which found *NFKB1* as a novel susceptible locus for SSc [19]. In addition, decrease in expression of *ARG1* in skin lesion of SSc with RNAP, contributing to suppression of *cellular response to transforming growth factor beta stimulus*, might be relevant to aberrant Th2 cell–M2 macrophage-mediated response, which is potent profibrotic mediators [17]. As to SSc with ATA, *cellular response to stress* was suppressed. Various cellular stresses are considered to trigger inflammation and immune signaling [20], suggesting they could be an initiator of aberrant response in skin lesion of SSc.

Fibrotic tissue responses are highly important aspects of SSc. The microvascular injury initiates a reparative cascade, triggering an inflammatory response and fibroblast activation [21]. In skin lesion of SSc with ACA, *keratinocyte differentiation* and *wound healing* were upregulated. When tissue is injury, myofibroblasts invade and repair injured tissues, following they undergo apoptosis [22]. However, in SSc, myofibroblasts are chronically activated, and therefore wound healing response and tissue remodeling prolong [23]. Considering the result of enrichment analysis, SSc with ACA polarize towards aberrant tissue remodeling with less contribution of inflammatory response.

Although patients without specific autoantibody were also included in the current study, the number of DEGs was disproportionally small. Except autoantibodies measured in the GENISOS cohort, SSc-associated autoantibodies are present at low frequencies. They include antibodies directed to endothelial cells, fibrillin-1, fibroblasts, against matrix metalloproteinases, the PDGF receptor and the angiotensin II type 1 receptor, and each of them is associated with different clinical characteristics [24]. Therefore, the reason why only 2 DEGs were generated from them because they might be heterogenous population.

This study suffers from several limitations. First, the possibility that factors other than autoantibody affected the result couldn’t be denied. If clinical information will be in available, confounding factor can be clear. Second, histological and functional experiments were needed to confirm that the dysregulated pathways contributed clinical phenotype.

In conclusion, distinct pathways were associated with type of autoantibody, while a part of gene expression profiling were overlapped among them, by leveraging gene expression data of patients and controls from multi-center cohort. The current study will promote to explore new therapeutic target for SSc.

## Data Availability

Supplementary Table 1, 2 and R script in the current study are available in figshare (https://figshare.com/articles/Supplementary_Table_1/12017049).

## Acknowledgement

Gene expression datasets were obtained from the Genetics versus Environment in Scleroderma Outcome Study (GENISOS) cohort. I thank all investigators for sharing the data.

## Data Availability

Supplementary Table 1,2 and R script in the current study are available in figshare (https://figshare.com/articles/Supplementary_Table_1/12017049).

## Authors’ Contributions

All of conceptualization, formal analysis and writing were conducted by the author.

## Ethics approval

Not required because this study uses only publicly available data.

## Consent for publication

Not required.

## Patient and Public Involvement

This research was done without patient involvement. Patients were not invited to comment on the study design and were not consulted to develop patient relevant outcomes or interpret the results. Patients were not invited to contribute to the writing or editing of this document for readability or accuracy.

## Figure legends

**Supplementary Table 1. Cluster of genes in hierarchical clustering analysis**.

**Supplementary Table 2. Genes with adjusted p-value less than 0**.**05 compared with healthy controls**.

ACA, anti-centromere antibody; ATA, anti-topoisomerase antibody; RNAP, anti-RNA polymerase 3 antibody; RNP, anti-U1RNP antibody.

